# Associations of Epigenetic Age Acceleration with Motor Impairment: Evidence from the PPMI Cohort

**DOI:** 10.64898/2025.12.16.25342400

**Authors:** Tao Jiang, Julia Hui, Bowen Feng, Guojun Tan, Guangwei Yuan, Jingyun Yang

**Author notes:** Correspondence to: Jingyun Yang, Rush Alzheimer’s Disease Center Rush University Medical Center, 1750 W Harrison ST, STE 1000, Chicago, IL 60612, USA.

## Abstract

**Objective:** Motor decline is a hallmark of Parkinson’s disease (PD) and biological aging. While epigenetic clocks measure systemic biological aging, their specific relationship with neuromotor function in the context of aging and neurodegeneration remains under-characterized. This study utilized data from the Parkinson’s Progression Markers Initiative (PPMI) to examine the relationships between multiple DNA methylation-based epigenetic aging measures and motor impairment across distinct domains.

**Methods:** We utilized longitudinal phenotype and whole-blood DNA methylation data from the PPMI cohort. Seven epigenetic aging measures were calculated, including first-generation clocks (Horvath v1, Hannum), risk-optimized second-generation clocks (PhenoAge, GrimAge v1 and v2), a deep-learning estimator (AltumAge), and a pace-of-aging biomarker (DunedinPACE). Motor function was assessed using the MDS-UPDRS, which captures patient-reported disability, global staging (Hoehn & Yahr), and specific motor signs, including tremor, bradykinesia, and postural instability. Cross-sectional associations were evaluated using binary or ordinal logistic regression, while longitudinal repeated measures were analyzed using generalized estimating equations (GEE) and cumulative link mixed models (CLMM), with adjustments for multiple comparisons made using the false discovery rate (FDR).

**Results:** Cross-sectional analyses yielded limited evidence, identifying only a specific association between Horvath v1 acceleration and postural tremor in males (P_FDR_ < 0.1). Repeated-measures analyses in the overall sample revealed robust associations between accelerated epigenetic aging and worsening tremor phenotypes (postural and kinetic), while associations with gait, rigidity, and bradykinesia were largely non-significant or inversely related. Sex-stratified analyses revealed distinct sexual dimorphism: males exhibited a broad, multi-clock phenotype (involving Horvath v1, PhenoAge, and GrimAge), where accelerated aging was associated with worsening tremor, gait, and rigidity. Females showed restricted associations, primarily linking the Hannum clock to tremor and global staging, and DunedinPACE to rigidity.

**Conclusions:** Accelerated epigenetic aging is robustly associated with the longitudinal progression of tremor impairment in PD, with associations being particularly consistent and multi-dimensional in men. These findings suggest that systemic biological aging, as captured by both first- and second-generation clocks, disproportionately exacerbates tremor circuitry distinct from the dopaminergic pathways that drive bradykinesia. Epigenetic clocks may serve as valuable biomarkers for monitoring progression and risk stratification, particularly for tremor-dominant phenotypes.

## Introduction

Aging is a complex biological process marked by a gradual decline in physiological integrity across multiple organ systems. This decline ultimately reduces resilience and increases vulnerability to disease and disability[1]. Impairments in motor function, such as slowed voluntary movement, diminished balance, alterations in gait, and reduced coordination, are among the most salient manifestations of aging[2, 3]. These motor impairments are driven by multifactorial mechanisms, including age-related neurodegeneration, sarcopenia, loss of proprioception, and impaired neuromuscular control[4, 5]. Motor decline is a core feature of biological aging. It exerts profound effects on quality of life, being closely linked with loss of independence, increased fall risk, and greater morbidity and mortality in older adults[6-8]. Therefore, understanding the biological mechanisms that contribute to motor deterioration with aging is crucial from both clinical and public health perspectives.

Epigenetic clocks, DNA methylation-based estimators of biological aging, quantify age-related physiological decline in a manner complementary to chronological age. First-generation clocks trained to predict chronological age include Horvath’s pan-tissue clock and the blood-derived Hannum clock, both of which introduced the now standard concept of “epigenetic age acceleration” (the difference between DNAm age and chronological age) as a person-level index of faster or slower biological aging[9, 10]. Subsequent “second-generation” designs optimized for health risk, such as DNAm PhenoAge and DNAm GrimAge, were trained on clinical phenotypes and mortality surrogates, respectively, and show robust associations with all-cause mortality, multimorbidity, and cardiometabolic disease; GrimAge version 2 further improves risk stratification by incorporating DNAm surrogates for CRP and HbA1c[11-13]. In parallel, rate-based measures, such as the DunedinPACE, assess the speed of biological aging and relate to morbidity, disability, and mortality across cohorts[14]. More recently, deep-learning clocks, such as AltumAge, have enhanced cross-tissue accuracy, underscoring that methylation captures multidimensional features of aging biology beyond linear CpG effects[15]. Importantly, faster epigenetic age acceleration has been linked not only to survival and chronic disease but also to aging-relevant functional phenotypes, including lower physical functioning and mobility (e.g., gait speed, grip strength) in both longitudinal and cross-sectional analyses, a greater frailty burden, and worse cognitive outcomes[11, 16-18]. These findings position epigenetic clocks and their acceleration metrics as integrative biomarkers that bridge molecular aging processes with clinically salient declines in healthspan and function.

Linking DNA methylation–based aging measures to motor function addresses a critical knowledge gap: do systemic biological aging processes, as registered by epigenetic clocks, map onto clinically salient decrements in neuromotor performance? Previous studies found that higher epigenetic age acceleration and faster rate-of-aging measures are associated with poorer physical functioning, such as slower gait speed, weaker grip, lower composite function, and greater frailty burden[14, 16-21]. Risk-optimized clocks (e.g., DNAm, GrimAge/GrimAge2) and rate-based biomarkers (e.g., DunedinPACE) are particularly informative, given robust links to disability and mortality and the incorporation of inflammatory and cardiometabolic surrogates that plausibly impinge on neuromuscular function[12-14]. In Parkinson’s disease (PD), epigenetic age acceleration has been linked to earlier age at onset. PD-related methylation signatures have been tied to motor progression, indicating that molecular aging signals may intersect with disease-specific motor decline[22, 23]. Accordingly, systematically comparing multiple clocks, including first-generation, risk-optimized, and rate-based measures, against distinct motor domains can identify which epigenetic metrics best index neuromotor aging and may aid risk stratification and progression monitoring in PD populations.

Therefore, in this study, we leveraged data from the Parkinson’s Progression Markers Initiative (PPMI) to test whether DNA–methylation–based measures of epigenetic aging are associated with motor function across multiple domains. Using seven widely used clocks, we evaluated links between biological aging signals and clinically salient motor phenotypes in Parkinson’s disease. Together, these analyses aim to identify epigenetic metrics that best capture neuromotor aging and inform risk stratification and monitoring.

## Methods

### Data sources

All phenotype and DNA-methylation data were obtained from PPMI, an international, longitudinal cohort launched in 2010 to accelerate biomarker discovery in Parkinson’s disease[24, 25]. PPMI enrolls three broad cohorts across multiple international sites under a harmonized protocol: recently diagnosed, untreated PD; healthy controls; and prodromal/genetic-risk groups. Participants undergo standardized clinical motor assessments, imaging, and biospecimen collection, with dense visits in year 1 and semiannual to annual follow-up thereafter[25]. The study emphasizes deep phenotyping and open data sharing through a centralized repository, from which we derived the analytic dataset for the present work. The analytic framework is illustrated in **Figure 1**.

**Figure 1.**
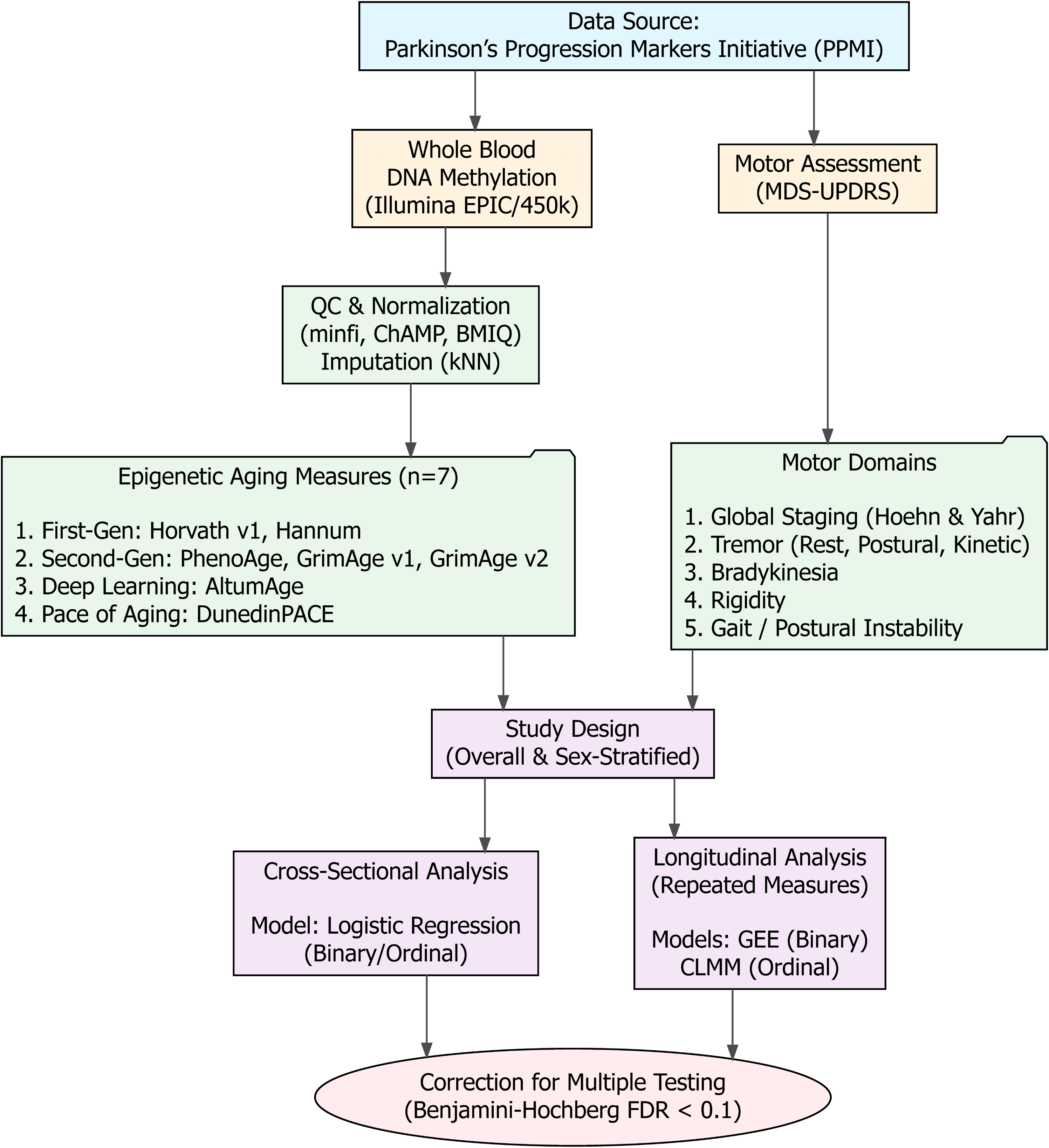
Schematic overview of the study design and analytical workflow. The study utilized longitudinal clinical and biological data from the Parkinson’s Progression Markers Initiative (PPMI). Raw DNA methylation data were processed to calculate seven epigenetic aging measures, while motor function was characterized across five domains using the MDS-UPDRS. Associations were evaluated using cross-sectional (logistic regression) and longitudinal (GEE/CLMM) models, stratified by sex, with correction for multiple testing. Abbreviations: BMIQ, Beta-Mixture Quantile normalization; CLMM, Cumulative Link Mixed Models; FDR, False Discovery Rate; GEE, Generalized Estimating Equations; kNN, k-Nearest Neighbors imputation; MDS-UPDRS, Movement Disorder Society-Sponsored Revision of the Unified Parkinson’s Disease Rating Scale; QC, Quality Control.

### Assessment of motor functions

PPMI assesses motor function with the MDS-UPDRS under a harmonized protocol and stores both patient-reported and examiner-rated measures. These assessments comprise four domains: 1) patient-reported motor disability; 2) global motor staging; 3) examiner-rated motor signs; and 4) motor complications. Per protocol, Part III examinations may be performed in defined OFF/ON states once dopaminergic therapy begins. In this study, to ensure that motor scores reflected endogenous motor decline rather than medication response, we utilized only data collected in the unmedicated or defined OFF state.

Assessments performed in the ON state or OFF-state assessments with undefined washout periods were excluded from the analysis. PPMI also captures binary clinical features of Parkinsonism (e.g., bradykinesia present, postural instability, and rest tremor present) at scheduled visits. Collectively, these instruments are well-validated and deeply phenotyped within PPMI, enabling domain-specific tests of neuromotor function[26]. For the complete list of items examined in this study, see **Supplementary Table 1**.

### DNA methylation data

Peripheral whole blood was collected at baseline and at planned follow-up visits following PPMI’s longitudinal protocol. DNA methylation profiling is released as raw Illumina Infinium array .idat files in the PPMI Methylation Profiling dataset (Projects 120/140). These arrays are predominantly HumanMethylationEPIC BeadChips. Repeated blood collections are an intentional feature of PPMI’s design, enabling within-person profiling of biological change. As a result, some participants contribute multiple DNA methylation assessments over time. Visit schedules (core PD and related cohorts) specify regular, repeated motor and biospecimen assessments, supporting the longitudinal acquisition of omics data.

We processed raw Illumina Infinium files in batches of roughly 500 samples using *minfi* and *ChAMP*[27, 28]. Only samples with both red and green channel files were retained. At the probe level, we removed any CpG that failed detection in any sample (detection p-value > 0.01) and probes supported by fewer than three beads in more than 5% of samples. To address missing or low-quality signals, we first excluded probes with >20% missingness and samples with >10% missingness, then imputed the remaining missing values using 5-nearest neighbors. We normalized β-values with BMIQ to correct type I/II probe bias[29] and mitigated technical batch effects with ComBat[30] before saving the resulting β-value matrices for analysis.

### Calculation of epigenetic clocks

Using the post-processed DNA methylation data, we calculated seven epigenetic clocks, including Horvath v1, Hannum, PhenoAge, GrimAge v1, GrimAge v2, DunedinPACE, and AltumAge. We selected this panel to cover complementary constructs and best-in-class performance: two first-generation chronological clocks (Horvath v1, Hannum), three second-generation morbidity/mortality-optimized clocks (PhenoAge, GrimAge v1/v2), a pace-of-aging biomarker with high test–retest reliability and strong prognostic validity (DunedinPACE), and a modern deep-learning pan-tissue estimator with improved cross-tissue age prediction (AltumAge). More detailed information regarding the seven clocks is provided in **Supplementary Table 2**.

The epigenetic aging measures were calculated primarily in R using the *meffonym* package, with the exception of AltumAge, which was implemented through the *pyaging* Python package. All calculations followed the developers’ recommended procedures, including optional calibration steps for Horvath and DunedinPACE, as well as the incorporation of chronological age and sex into GrimAge estimates to ensure consistency with the original algorithms. Epigenetic age acceleration was defined as the difference between epigenetic age and chronological age, with positive values indicating faster biological aging. For DunedinPACE, no subtraction was necessary, as the measure itself directly reflects the pace of aging.

### Statistical analysis

We first summarized participant characteristics using descriptive statistics. Categorical variables were reported as counts (percentages), and continuous variables as means with standard deviations.

We conducted cross-sectional analyses by utilizing data from the first available motor assessment for each outcome per participant. Associations between each epigenetic clock and motor outcomes were evaluated using binary or ordinal logistic regression models, as appropriate, adjusting for sex. To fully leverage multiple assessments of motor functions, we then conducted longitudinal analyses by incorporating all available repeated motor assessments. Binary motor outcomes (e.g., presence of bradykinesia, postural instability, or rest tremor) were modeled using generalized estimating equations (GEE), which account for within-subject correlation by specifying a working correlation structure and provide population-averaged estimates that are robust to misspecification of the correlation matrix[31]. Ordinal outcomes (e.g., Hoehn & Yahr stage, UPDRS ordinal items) were modeled using cumulative link mixed models (CLMM), which extend proportional odds models by including subject-specific random effects to account for repeated measures and intra-individual correlation. Both GEE and CLMM models included sex as a covariate.

For both cross-sectional and longitudinal analyses, given the well-documented sexual dimorphism in Parkinson’s disease phenotypes and the known differences in epigenetic aging between sexes, we performed pre-planned analyses stratified by sex to examine potential subgroup patterns. All tests were two-sided, with P < 0.05 considered statistically significant. To address the issue of multiple hypothesis testing across the seven clocks and multiple motor outcomes, we calculated the Benjamini-Hochberg False Discovery Rate (FDR). We considered associations with an FDR-adjusted q-value < 0.10 to be robust, though we prioritized reporting effect sizes and patterns of association consistent with biological hypotheses. All statistical analyses were performed using R version 4.4.2.

## Results

**Supplementary Table 3** provided the basic characteristics of the study participants across motor outcomes. Across motor outcomes, the number of participants with non-missing data ranged from 216 to 515. The mean age at the visit was ∼64 years (SD ∼9), and about 60% of them were male. The majority of them were White, with a mean education of around 16 years (SD ∼3.6). Epigenetic aging metrics showed roughly similar distributions across outcomes.

Participants displayed heterogeneity across the major motor domains. Binary clinical features showed that bradykinesia was the most common (62%), followed by rest tremors (45%) and postural instability (43%). Global motor staging by Hoehn & Yahr indicated that most participants were in early disease stages: 20% at stage 1, 70% at stage 2, and fewer than 10% at stage 3 or above. Examiner-rated signs from UPDRS Part III showed notable prevalence of motor impairment, including gait disturbance, freezing of gait, and tremor (rest, kinetic, and postural), and rigidity affecting the neck, upper, and lower extremities. More detailed information on the distribution of these outcomes is provided in **Supplementary Table 4**.

In the cross-sectional analysis of the overall sample (**Supplementary Table 5**), we found no robust evidence that accelerated epigenetic aging was associated with worse motor outcomes (OR > 1) after FDR correction. Some first-generation measures (Horvath v1, Hannum) and AltumAge showed significant associations with global staging (Hoehn & Yahr) and postural instability (P_FDR_ < 0.1). However, these associations were consistently in the inverse direction (OR < 1), suggesting potential confounding or selection effects in baseline cross-sectional comparisons.

Sex-stratified cross-sectional analyses yielded similarly sparse results. In females, no associations survived FDR correction (**Supplementary Table 6**). In males (**Supplementary Table 7**; **Figure 2**), we observed a specific signal for left-sided postural tremor: Horvath v1 acceleration was associated with greater odds of postural tremor (OR = 1.14, 95% CI 1.06–1.24, P = 0.0007, P_FDR_ = 0.08). Several other accelerated aging measures, including PhenoAge, GrimAge v1, GrimAge v2, and AltumAge, showed nominal associations with the same outcome; however, these did not survive FDR correction.

**Figure 2.**
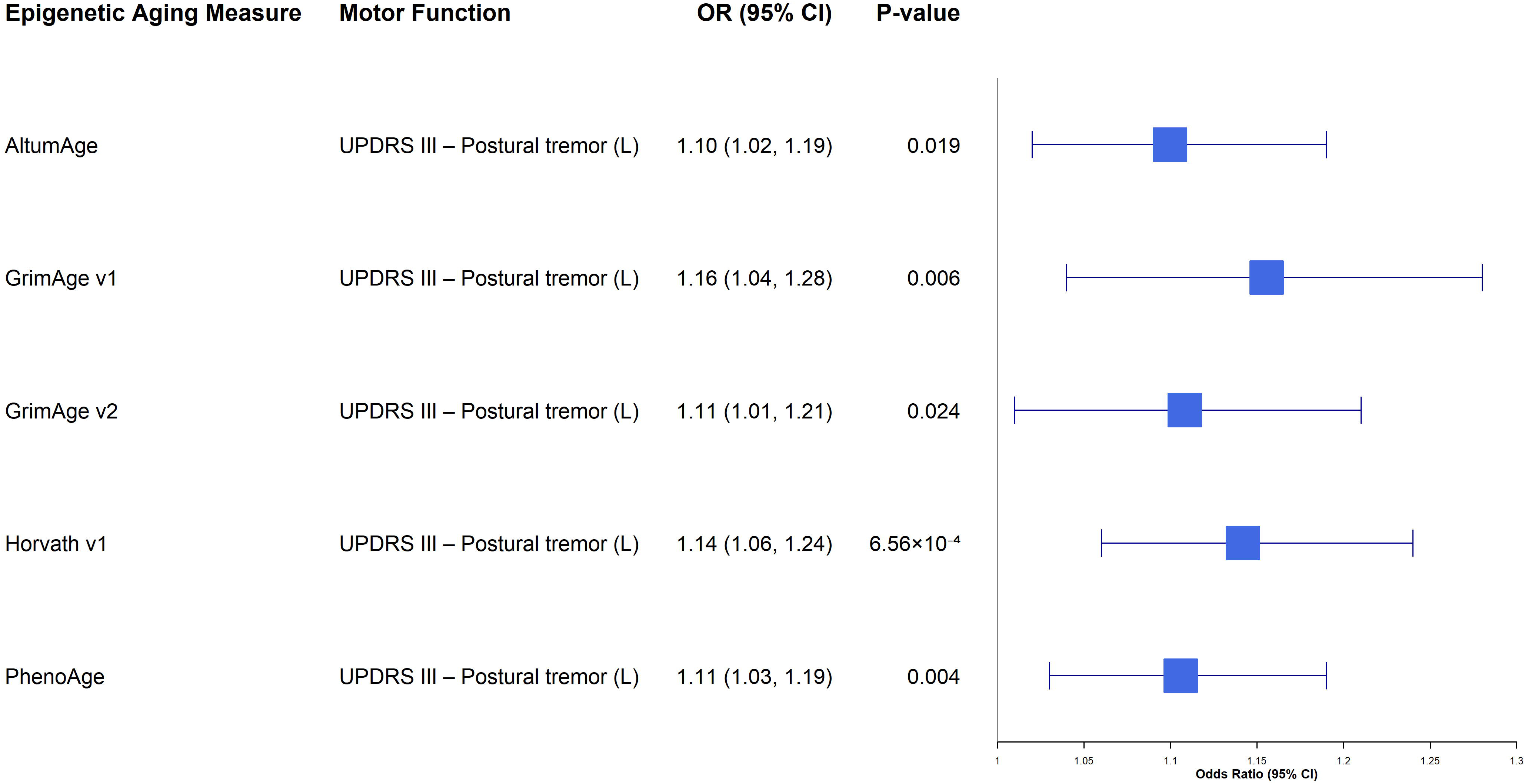
Forest plot of cross-sectional associations between epigenetic age acceleration and motor function in males. This plot displays the associations between epigenetic aging measures and motor outcomes in the male cohort using baseline data. To focus on biologically relevant risk signals, only associations with a nominal p-value < 0.05 and odds ratio (OR) > 1 (indicating that accelerated aging is associated with worse motor function) are presented. The x-axis represents the odds ratio (OR) on a linear scale, and error bars indicate the 95% confidence intervals. Abbreviations: CI, confidence interval; OR, odds ratio.

Longitudinal analyses incorporating repeated measures in the overall sample (**Supplementary Table 8**; **Figure 3**) revealed a coherent signal linking accelerated aging to worsening tremor phenotypes. For left-sided postural tremor, GrimAge v2 (OR = 1.05, 95% CI 1.05–1.06, P_FDR_ < 0.001) and DunedinPACE (OR = 1.02, 95% CI 1.01–1.02, P_FDR_ < 0.001) showed robust associations, while Hannum (OR = 1.02, 95% CI 1.02–1.03, P_FDR_ < 0.001) was associated with right-sided postural tremor. Additionally, Hannum and GrimAge v1 showed nominal associations with left-sided postural tremor (P < 0.05), though these did not pass FDR correction. GrimAge v1 acceleration was also associated with increased odds of left-sided kinetic tremor (OR = 1.08, 95% CI 1.07–1.08, P_FDR_ < 0.001). In contrast, associations with gait, rigidity, and bradykinesia in the overall sample were generally weaker, inconsistent, or in the inverse direction, indicating domain specificity of the aging signal.

**Figure 3.**
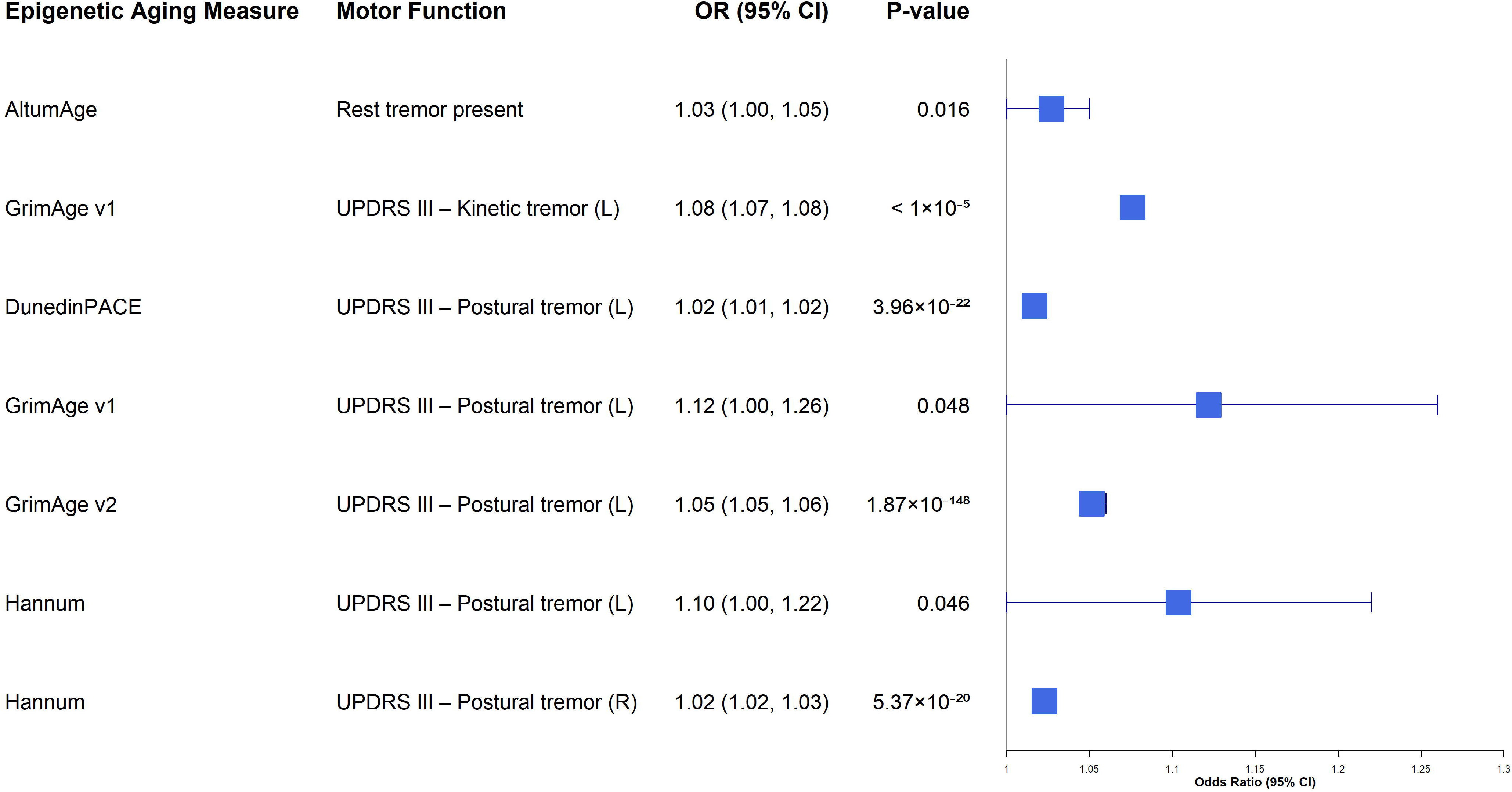
Forest plot of longitudinal associations between epigenetic age acceleration and motor function in the overall sample. This plot illustrates the results from repeated-measures analyses (using generalized estimating equations or cumulative link mixed models) in the full analytic cohort. To emphasize risk-consistent signals, only associations with nominal P < 0.05 and OR > 1 are displayed. Error bars denote 95% confidence intervals. Abbreviations: CI, confidence interval; OR, odds ratio; PD, Parkinson’s disease.

Sex-stratified longitudinal models revealed that the motor domains associated with biological aging differed by sex (**Supplementary Tables 9 & 10**). In females (**Supplementary Table 9**; **Figure 4)**, robust associations in the expected direction were restricted, with Hannum acceleration related to worse global staging (Hoehn & Yahr Stage, OR = 1.08, P_FDR_ < 0.001) and left-sided postural tremor (OR = 1.03, P_FDR_ < 0.001), and DunedinPACE associated with increased rigidity in the left upper extremity (OR = 1.05, P_FDR_ < 0.001). In males (**Supplementary Table 10**; **Figure 5**), we observed broader multi-clock associations centered on tremor and gait involving both first- and second-generation clocks. Horvath v1 (OR = 1.22, P_FDR_ = 0.006), PhenoAge (OR = 1.15, P_FDR_ = 0.038), and GrimAge v1 (OR = 1.25, P_FDR_ = 0.066) were associated with greater odds of left-sided postural tremor, while PhenoAge (OR = 1.04, P_FDR_ < 0.001) and GrimAge v2 (OR = 1.02, P_FDR_ < 0.001) were associated with left-sided kinetic tremor. In addition, PhenoAge acceleration was associated with worse Gait scores (OR = 1.01, P_FDR_ < 0.001), and Hannum acceleration was associated with increased rigidity in the left upper extremity (OR = 1.02, P_FDR_ < 0.001).

**Figure 4.**
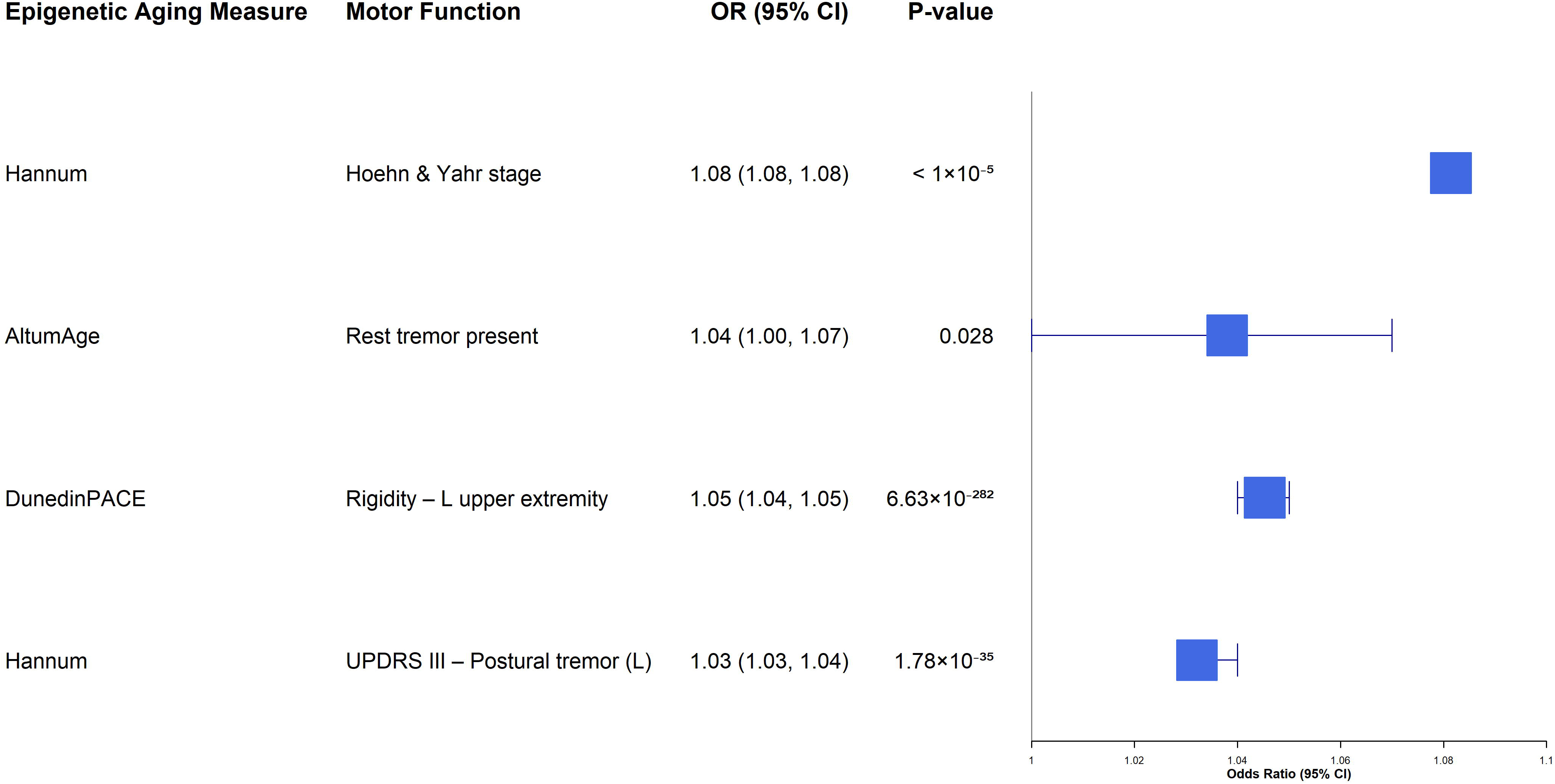
Forest plot of longitudinal associations between epigenetic age acceleration and motor function in females. This plot presents the results from sex-stratified repeated-measures analyses restricted to females. Only associations with nominal P<0.05 and OR > 1 are displayed. Error bars denote 95% confidence intervals. Abbreviations: CI, confidence interval; OR, odds ratio.

**Figure 5.**
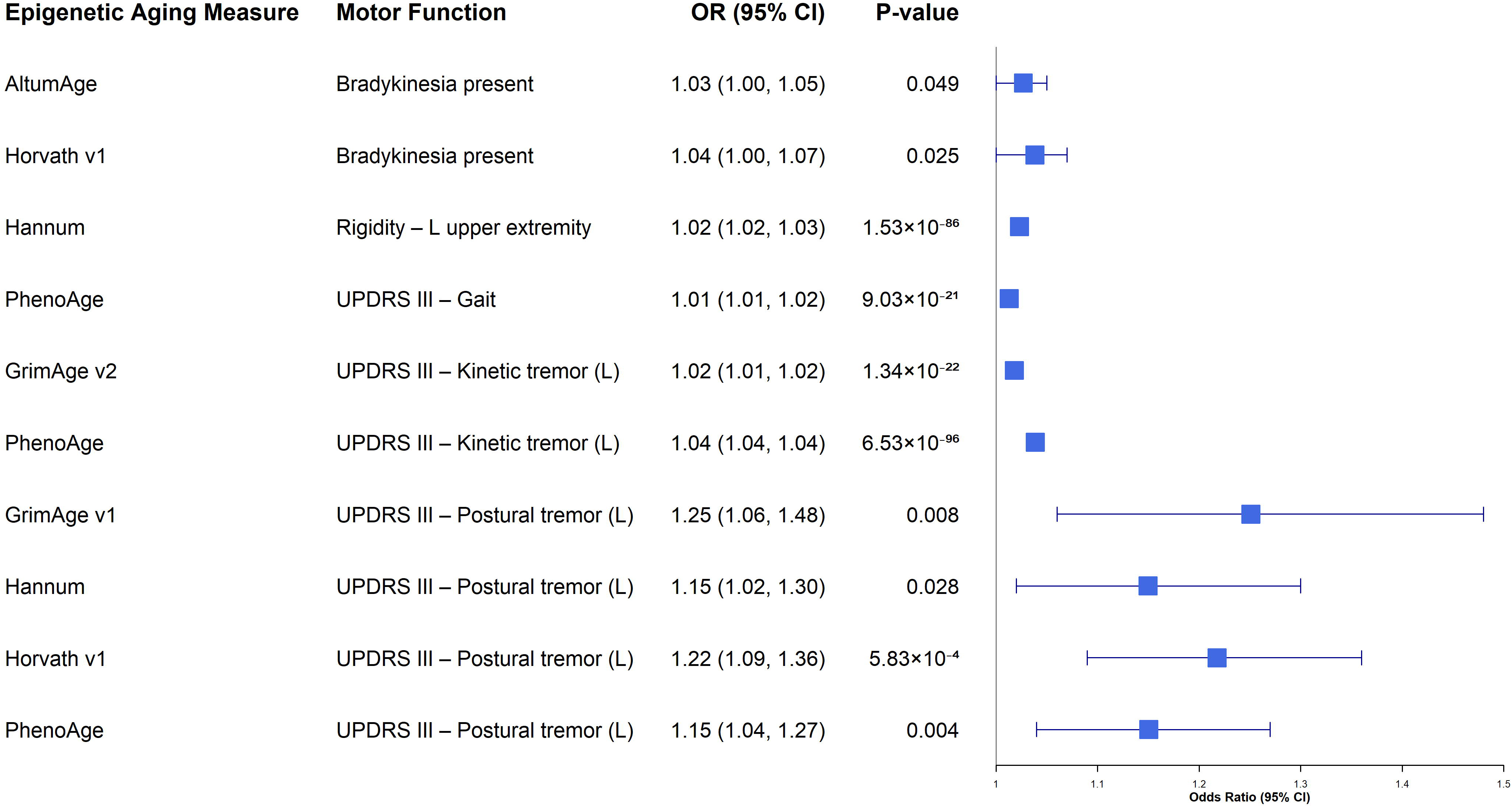
Forest plot of longitudinal associations between epigenetic age acceleration and motor function in males. This plot presents the results from sex-stratified repeated-measures analyses restricted to males. Only associations with nominal P<0.05 and OR > 1 are displayed. Error bars denote 95% confidence intervals. Abbreviations: CI, confidence interval; OR, odds ratio.

## Discussion

In this study, we utilized data from the PPMI cohort to examine the relationship between systemic biological aging and specific motor domains. Our primary finding is that epigenetic age acceleration is robustly and differentially associated with tremor phenotypes, including postural and kinetic tremors, while associations with rigidity and bradykinesia were largely inconsistent or inversely related. This "tremor-specific" signal was observed across multiple epigenetic measures, indicating that both first-generation (e.g., Hannum, Horvath v1) and risk-optimized (e.g., PhenoAge, GrimAge, and DunedinPACE) clocks capture biological processes relevant to tremor progression.

Furthermore, we identified distinct sex-specific profiles: while accelerated aging in females was primarily linked to tremor via the Hannum clock, males exhibited a pervasive multi-clock phenotype where accelerated aging tracked with worsening tremor, gait, and global staging. These findings suggest that systemic biological aging may exacerbate tremor circuitry and postural control mechanisms, distinct from the dopaminergic pathways driving bradykinesia.

A major finding of this study is the domain-specific association between accelerated epigenetic aging and tremor phenotypes (postural and kinetic), in contrast to the null or inverse associations observed with rigidity and bradykinesia. This divergence likely reflects the distinct pathophysiological mechanisms underlying these motor domains. While bradykinesia and rigidity are the canonical consequences of focal nigrostriatal dopamine depletion and respond robustly to dopaminergic therapy [32, 33], parkinsonian tremor, particularly postural and kinetic forms, involves a more widespread, distinct neural network comprising the cerebello-thalamo-cortical loops[34, 35]. Our results suggest that systemic biological aging, as captured by epigenetic clocks, may disproportionately exacerbate this distributed "tremor network," whereas rigidity remains primarily driven by disease-specific a-synuclein pathology that operates independently of background aging rates[36]. This hypothesis aligns with evidence that epigenetic age acceleration in older adults reflects "systemic integrity" and physiological frailty rather than isolated organ dysfunction[16, 37]. In the context of PD, patients with accelerated biological aging may experience a compounded decline where systemic neuronal vulnerability amplifies widespread tremor circuitry, while the focal dopaminergic drivers of bradykinesia remain less sensitive to these systemic aging processes. Consequently, epigenetic clocks may serve as specific biomarkers for the non-dopaminergic or age-dependent components of motor progression, characterizing a "tremor-dominant" trajectory distinct from the akinetic-rigid phenotype[38].

Through our longitudinal analyses, we found that men exhibited a broad, multi-clock association with motor decline while women showed restricted associations. This sexual dimorphism parallels established biological sex differences in both aging and Parkinson’s disease. In the general population, males consistently exhibit accelerated epigenetic aging rates in blood and brain tissue compared to age-matched females, a phenomenon known as the "male disadvantage" in biological aging[39, 40]. Our findings suggest this accelerated baseline velocity in men may translate into a tighter coupling between systemic aging and the degradation of tremor and postural control mechanisms, effectively lowering the "frailty threshold" at which cellular damage manifests as clinical symptoms. Conversely, the attenuated signal in the female cohort aligns with the neuroprotective "estrogen reserve" hypothesis, which posits that estrogen modulates dopaminergic neurotransmission and mitigates oxidative stress, potentially buffering the neural networks governing tremor against systemic aging pressures[41, 42].

Consequently, women may require more specific or severe biological perturbations, such as those captured by the Hannum clock, which is highly sensitive to blood-based immune senescence[10], to override this protective reserve and drive tremor progression. This divergence highlights the importance of considering sex as a fundamental biological variable in biomarker development. Epigenetic clocks may possess higher prognostic utility for tremor-dominant phenotypes in men[43], whereas female-specific risk stratification may require markers that account for hormonal and immune-mediated resilience.

This study has several limitations that should be considered. First, the sample size was limited, particularly in the sex-stratified analyses. This reduces statistical power and likely contributes to the attenuated or restricted findings in the female cohort. Consequently, the lack of multi-clock associations in women should be interpreted with caution, and we cannot rule out that broader effects exist but were undetectable in this sample; future studies with larger cohorts are needed to validate the female-specific signal observed with the Hannum clock.

Second, our results highlight the inherent limitations of cross-sectional "snapshots" in Parkinson’s disease research. We observed that baseline cross-sectional analyses yielded largely null or inverse associations, likely due to the profound heterogeneity of PD motor phenotypes at study entry[33]. In contrast, our repeated-measures analyses successfully captured the trajectory of decline, underscoring that biological aging markers track the rate of progression rather than static disease severity. However, while we incorporated repeated measures, the number of follow-up assessments was modest, potentially preventing the full capture of long-term epigenetic trajectories.

Third, our analysis was conducted predominantly among White participants. Our findings may not be generalized to more diverse populations where epigenetic aging patterns and Parkinson’s disease features may differ. In addition, all methylation measures were derived from peripheral blood, which may not fully reflect tissue-specific aging processes in the central nervous system, although prior work indicates that blood-based clocks effectively capture systemic inflammatory and metabolic pathways relevant to neurodegeneration[11].

Another consideration is the multiplicity of statistical testing. We addressed multiple hypothesis testing across clocks and outcomes using the Benjamini–Hochberg false discovery rate; however, some nominal associations may still reflect chance findings, given the correlated outcomes and aging measures. Finally, although we restricted our analyses to unmedicated or defined OFF-state motor assessments, residual confounding by unmeasured factors, such as physical activity, comorbidities, or cumulative medication exposure, cannot be ruled out.

In conclusion, we found that accelerated epigenetic aging is robustly associated with the longitudinal progression of tremor and gait impairment, with a striking male-specific predisposition. These findings suggest that epigenetic clocks, including both risk-optimized measures such as GrimAge/PhenoAge and first-generation estimators such as Hannum, capture biological processes that disproportionately exacerbate tremor circuitry. Our results challenge the utility of cross-sectional biological aging markers in PD, advocating instead for their use as longitudinal biomarkers to monitor trajectory and aid in risk stratification, particularly for tremor-dominant phenotypes in men.

## Author Contributions

**Tao Jiang:** Conceptualization, Writing – original draft, Writing – review and editing

**Julia Hui:** Data curation, Formal analysis, Writing – original draft

**Bowen Feng:** Formal analysis, Visualization, Writing – original draft

**Guangwei Yuan:** Formal analysis, Visualization

**Jingyun Yang:** Conceptualization, Supervision, Writing – original draft, Writing – review and editing

## Ethics approval

This study used only publicly available data. As a result, ethical approval and consent to participate are not needed.

## Data Availability

The PPMI data generated or analyzed during this study are publicly available and can be requested at https://ida.loni.usc.edu/.

## Consent to participate

NA.

## Consent to publish

NA.

## Declaration of generative AI and AI-assisted technologies in the writing process

During the preparation of this work, the author(s) used ChatGPT (OpenAI)/Gemini to improve the clarity and readability of the language. After that, the author(s) carefully reviewed and edited the content to ensure accuracy and appropriateness and take full responsibility for the content of the published article.

